# Association between Binge Drinking Behaviors and Comorbidities in Brazil: Network Analysis of a National Health Survey

**DOI:** 10.1101/2023.12.18.23300120

**Authors:** Siddhesh Zadey, Diego Franca, Pollyana Coelho Pessoa Santos, Natan David Pereira, Yolande Pokam Tchuisseu, Luciano Andrade, Bruno Pereira Nunes, Wagner De Lara Machado, Catherine A. Staton, Joao Ricardo Nickenig Vissoci

**Affiliations:** Global Emergency Medicine Innovation and Implementation Research Center, Duke University, Durham North Carolina USA; Duke Global Health Institute, Durham North Carolina USA; Association for Socially Applicable Research (ASAR), Pune, Maharashtra, India; Dr. D.Y. Patil Medical College, Hospital, and Research Center, Pune, Maharashtra, India; State University of Maringá, Paraná, Brazil; Department of Nursing, State University of the West of Parana, Foz do Iguaçu, Parana, Brazil; Faculty of Nursing, Federal University of Pelotas, Pelotas, Rio Grande do Sul, Brazil; Pontifical Catholic University of Rio Grande do Sul, Porto Alegre, Brazil; Department of Emergency Medicine, Duke University School of Medicine, Durham North Carolina USA

## Abstract

Alcohol consumption is the sixth leading cause of death globally. Brazil ranks second in alcohol-related mortality within the American regions with a notable increase in binge drinking behavior from 2013 (5.9%) to 2019 (17.1%). Binge drinking, a form of alcohol misuse, is a known risk factor for several diseases. We aimed to understand the differences in binge drinking across various sociodemographic groups and the patterns of comorbidities in a national-level dataset by doing a cross-sectional network analysis of the 2013 Brazilian National Health Survey. Binge drinking was defined as a binary variable based on alcohol consumption of >5 (4) doses in a month for male (female) responders. Weighted undirected network analysis using Ising models was performed to discover the strength of inter-relations between nineteen chronic conditions. In the network, the nodes represented the conditions and the edges were formed by statistical associations derived using logistic regression. Community analyses identified the clusters within networks. A nationally representative sample of 60,202 people revealed the prevalence of binge drinking to be about 13.5%. The study revealed a less connected network among binge drinkers, potentially impacting disease associations. Binge drinking demonstrated unique correlations with comorbidities across age, gender, ethnicity, and education levels, highlighting the complex interplay between binge drinking and health outcomes. Recognizing the specific comorbidities associated with binge drinking, such as hypertension and chronic spinal problems, allows healthcare professionals to tailor preventive measures and early interventions. In this sense, differences in binge drinking and its direct association with comorbidities as well as in comorbidity structures across sociodemographic characteristics point to the utility of network models to identify specific populations at various health risks.

## Introduction

Alcohol consumption poses a significant global public health concern, with detrimental effects on individuals and societies worldwide. In 2018, the average alcohol consumption per person globally was 6.2 liters of pure alcohol (1), resulting in approximately 3 million deaths annually attributed to harmful alcohol use(2). Compared to American regions, Brazil ranks second in terms of alcohol consumed per person and second in alcohol-related mortality within the American regions, while having the seventh-highest heavy drinking prevalence in 2019(3,4). Furthermore, Brazil not only demonstrates high average alcohol consumption per person-year (7.67 liters)(5) but also a notable increase in binge drinking behavior from 2013 (5.9%) to 2019 (17.1%) (6).

The impact of alcohol consumption on public health is represented for more than 200 diseases alcohol-related, being the leading risk factor for disease burden among people aged 25 to 49, and the second leading risk factor among people 10 to 24 years old (1,7). The correlation between alcohol consumption and multimorbidities is well-established, with the risk of developing diseases being directly correlated to the quantity of alcohol consumed (8). As such, alcohol use serves as a risk factor for systemic hypertension, ischemic heart disease, ischemic stroke, and hepatic conditions, among others (8). The consumption of five or more drinks for men and four or more drinks for women on a single occasion, or binge drinking(9), further exacerbates the burden of diseases. For instance, melanoma, head and neck, testicular, and cervical cancer are more likely in people who report binge drinking behavior (10). A Brazilian study found higher odds of currently suffering from depression among the binge-drinking population aged 18-39 years old, and this finding was significantly greater in female binge drinkers (11).

Network analysis is a useful technique to investigate the patterns of associations among different conditions at a population scale that measures the strength of the multivariate correlations between several diseases (12). While network analysis has been employed to study comorbidity associations in specific contexts, such as the elderly or individuals with cardiovascular diseases (12,13), limited research exists on the application of network analysis to binge-drinking populations. Most published studies on this topic primarily explore its association with risky behaviors, environmental factors(14–17), or its correlation with major depressive disorders (18).

There is a paucity of network analysis literature on binge drinking and multimorbidity, particularly within a country with a high prevalence of alcohol consumption. As such, this study aims to examine the complex patterns of comorbidities and binge drinking behavior. We seek to compare the different multimorbidities correlations among binge drinking and non-binge drinking populations using the Brazilian National Healthy Survey. In addition, we aim to explore the interaction of binge drinking and morbidities within different demographic groups of that population.

## Materials and Methods

### Study design and Ethics

This was a retrospective secondary data analysis of a nationally representative cross-sectional survey. This national health survey has been de-identified and made publicly available, and as such an ethics or regulatory approval is not required according to Brazilian national policies.

### Setting & Study Population

The data was collected from the 2013 Brazilian National Health Survey (PNS, *Plano Nacional de Saúde*) conducted by the Brazilian Institute of Geography and Statistics (IBGE) (19). The survey encompassed a nationwide demographic representation by interviewing 62,986 households across all regions in Brazil, including both urban and rural areas, between January 2013 and December 2013. To gather information about family and household conditions, a single participant above the age of 14, who is deemed capable of providing the requested details, was interviewed in every household. The participants were asked about health access, use of health services, and lifestyle.

Each participant completed socio-demographic questions of the PNS questionnaire with information about sex (male, female), age (18 to 24 years, 25 to 44 years, 45 to 64 years, 65 years or older), race/ethnicity (white, black/parda, yellow-skinned/indigenous), region (North, Northeast, Midwest, Southeast, South), and education level (no schooling, elementary/equivalent, secondary/equivalent, post-secondary/equivalent).

### Chronic conditions

In this study, we selected 13 chronic conditions from the Q module of the PNS questionnaire: hypertension, diabetes, cardiac diseases (including myocardial infarction, angina, or congestive heart failure), stroke, asthma, arthritis/rheumatoid arthritis, chronic musculoskeletal back disorders, work-related musculoskeletal disorder (WMSD), depression, psychiatric conditions (including schizophrenia, bipolar disorder, or obsessive-compulsive disorder), pulmonary disorders (including chronic obstructive pulmonary disorder, chronic bronchitis or emphysema), cancer, and chronic kidney failure (19). For chronic musculoskeletal back disorders, the questionnaire asked “Do you have any chronic spinal problems, such as chronic back or neck pain, low back pain, sciatic pain, or vertebral or disc problems?” On the other hand, the other conditions were asked as follows: “Has a doctor ever given you the diagnosis of […]?” The 13 chronic conditions were dichotomously coded as no (0) or yes (1).

### Outcome

The PNS questionnaire measured binge drinking behavior using question 32 of module P, which asked “In the last 30 days, did you consume 5 or more doses of alcoholic beverage on one occasion, if male?” or “In the last 30 days, did you consume 4 or more doses of alcoholic beverage on one occasion, if female?” The answer options were no (0) and yes (1)(19).

### Statistical methods

#### Descriptive statistics

We reported counts and proportions for sociodemographic variables. Also, we estimated the overall prevalence of the chronic conditions, as well as their prevalence in the binge drinking context. Prevalence of the chronic conditions by sociodemographic variables is available in Supplementary Materials. We adjusted all the proportion/prevalence estimates by sample weights.

#### Network analysis

Network analysis is a popular technique to investigate associations among multiple recurrent diseases in a population. The use of network analysis in the investigation of behavioral associations within large populations is complex, but its application allows the identification of multiple connections between modifiable and non-modifiable behaviors and their role in developing diseases including multimorbidities. The visualization of these associations can be made by network maps, which demonstrate the nature, relationship intensity, directionality, and dimension of the variable correlations(20). In the context of multimorbidities, network analyses are able to measure the strength of associations of health conditions, representing them in a multivariate ’map’. Thus, network analysis points to the existence of the relationship among two or more morbidities and, more importantly, the association that the condition exerts on the other nodes within the network (12).

In this study, we performed seven sets of network models. The first set contains an overall network, with all 13 chronic conditions and binge drinking behavior as nodes. The second one comprised chronic conditions networks divided by binge drinking behavior. The other five sets have chronic conditions and binge drinking behavior as nodes and were divided by the categories of sex, age, race/ethnicity, region, and education level (Table 1). A network model is a statistical and visualization technique that allows one to explore the relationship between variables(21,22). In this context, ‘nodes’ represent variables, and ‘edges’ represent the conditional association between nodes. Blue (red) edges show positive (negative) conditional associations and the edge thickness reveals the weight of the association.

**Table 1:** Overview of the seven network sets.

| Network set | By | Chronic conditions | Binge drinking behavior |
| --- | --- | --- | --- |
| 1 | – | All | Yes |
| 2 | Binge drinking behavior | All | – |
| 3 | Sex | All | Yes |
| 4 | Age | All | Yes |
| 5 | Race/ethnicity | All | Yes |
| 6 | Region | All | Yes |
| 7 | Education level | All | Yes |

Due to the dichotomous nature of the variables, we estimated Ising Models for each network set, which performs nodewise logistic regressions(23). To handle false-positive coefficients, we used the Least Absolute Shrinkage and Selection Operator (LASSO)(24). LASSO penalizes all edge-weights toward zero and sets small weights to exactly zero, returning a sparser network. To choose the most appropriate strength of the penalty, we used the Extended Bayesian Information Criterion (EBIC) and set its tuning parameter *λ* to 0.25 (25). We computed centrality measures of betweenness (i.e., how often a node serves as the shortest path between other nodes), closeness (i.e., how close a node is, in average, to all other nodes), and expected influence (i.e., sum of all edge-weights connected to a node, considering the direction of the association), to investigate the importance of each node to the network structure (26,27).

We estimated node *predictability* for each node in order to measure how nodes connected to a particular node can predict it. In the network, predictability can be visualized by the ring around each node: colored areas mean the proportion of predictability each node has. We used the marginal correct classification (CCmarg) predictability measure. The interpretation is similar to the *R*^2^(28). To find node communities, we performed the *spinglass* algorithm, which implies that connected nodes should belong to the same community, and nodes of different communities should not present links between them (29). To check the edge-weight accuracy and the centrality measures stability, we estimated 1000 new samples through *bootstrapping* (23). Afterwards, we calculated bootstrapped confidence intervals for edge-weights and correlation stability coefficients (CS-coefficient) for centrality measures.

Data analysis was conducted in R software (v. 4.1.1) (30). We used the following R packages: *survey* (31) to adjust prevalences/proportions by sample weights, *mgm* (32) to estimate networks, *qgraph* (33) to visualize the networks and compute centrality measures, *igraph* (34) to find node communities, and *bootnet* (23) to perform bootstrapping analysis.

## Results

This study analyzed a total of 60,202 individuals, including 8,104 people with binge drinking behavior and 52,098 with no binge drinking behavior. In both analyzed groups, the majority of the population was from the Southeast region, reported to be black, and aged between 25 to 44 years old (Table 2). Among the binge drinking group, males presented the majority (74.5%), compared to females (25.5%), and 37.7% of the total binge drinking population completed a secondary educational level. In addition, the Northeast and Midwest regions presented a higher prevalence of people in the binge drinking group. People with hypertension and chronic spinal problems were the two groups with the highest prevalence of binge drinking, 13.7%, and 14.7% respectively (Table 3).

**Table 2:** Study Sample characteristics by binge drinking behavior status.

| Characteristic | Overall |  | Binge drinking behavior |  | No binge drinking behavior |  |
| --- | --- | --- | --- | --- | --- | --- |
|  | N = 60,202<sup>1</sup> | % (95% CI)<sup>2</sup> | N = 8,104<sup>1</sup> | % (95% CI)<sup>2</sup> | N = 52,098<sup>1</sup> | % (95% CI)<sup>2</sup> |
| **Sex** |  |  |  |  |  |  |
| Male | 25,920 | 47.1% (46.4%, 47.9%) | 5,702 | 74.5% (72.8%, 76.1%) | 20,218 | 42.8% (42.0%, 43.6%) |
| Female | 34,282 | 52.9% (52.1%, 53.6%) | 2,402 | 25.5% (23.9%, 27.2%) | 31,880 | 57.2% (56.4%, 58.0%) |
| **Age groups (years)** |  |  |  |  |  |  |
| 18 to 24 | 7,823 | 15.9% (15.4%, 16.5%) | 1,239 | 20.1% (18.4%, 22.0%) | 6,584 | 15.3% (14.7%, 15.9%) |
| 25 to 44 | 26,740 | 40.8% (40.1%, 41.6%) | 4,612 | 53.0% (51.1%, 54.9%) | 22,128 | 38.9% (38.1%, 39.7%) |
| 45 to 64 | 17,927 | 31.0% (30.3%, 31.7%) | 2,017 | 24.3% (22.8%, 25.9%) | 15,910 | 32.0% (31.3%, 32.8%) |
| 65 or older | 7,712 | 12.3% (11.8%, 12.8%) | 236 | 2.6% (2.1%, 3.2%) | 7,476 | 13.8% (13.3%, 14.4%) |
| **Race** |  |  |  |  |  |  |
| White | 24,106 | 47.5% (46.7%, 48.3%) | 2,887 | 43.2% (41.3%, 45.1%) | 21,219 | 48.1% (47.3%, 49.0%) |
|  | N = 60,202<sup>1</sup> | % (95% CI)<sup>2</sup> | N = 8,104<sup>1</sup> | % (95% CI)<sup>2</sup> | N = 52,098<sup>1</sup> | % (95% CI)<sup>2</sup> |
| Black/Parda | 35,146 | 51.2% (50.4%, 52.0%) | 5,082 | 55.6% (53.7%, 57.5%) | 30,064 | 50.5% (49.6%, 51.3%) |
| Yellow-skinned/Indigenous | 950 | 1.4% (1.2%, 1.5%) | 135 | 1.2% (0.9%, 1.5%) | 815 | 1.4% (1.2%, 1.6%) |
| **Region** |  |  |  |  |  |  |
| North | 12,536 | 7.4% (7.2%, 7.6%) | 1,657 | 7.7% (7.0%, 8.5%) | 10,879 | 7.4% (7.2%, 7.6%) |
| Northeast | 18,305 | 26.6% (26.1%, 27.1%) | 2,746 | 30.4% (28.8%, 32.2%) | 15,559 | 26.0% (25.5%, 26.6%) |
| Midwest | 7,519 | 7.4% (7.2%, 7.6%) | 1,144 | 8.7% (8.0%, 9.5%) | 6,375 | 7.2% (6.9%, 7.4%) |
| Southeast | 14,294 | 43.8% (43.1%, 44.4%) | 1,778 | 41.1% (39.0%, 43.2%) | 12,516 | 44.2% (43.5%, 44.9%) |
| South | 7,548 | 14.8% (14.4%, 15.2%) | 779 | 12.0% (10.8%, 13.3%) | 6,769 | 15.2% (14.7%, 15.7%) |
| **Education level** |  |  |  |  |  |  |
| No schooling | 9,434 | 13.7% (13.2%, 14.2%) | 845 | 8.8% (7.8%, 9.9%) | 8,589 | 14.5% (13.9%, 15.1%) |
| Elementary/equivalent | 20,537 | 35.2% (34.4%, 35.9%) | 2,728 | 33.7% (31.9%, 35.7%) | 17,809 | 35.4% (34.6%, 36.2%) |
| Secondary/equivalent | 19,438 | 33.6% (32.9%, 34.4%) | 2,973 | 37.7% (35.7%, 39.8%) | 16,465 | 33.0% (32.2%, 33.8%) |
| Post-secondary/equivalent | 10,793 | 17.5% (16.7%, 18.3%) | 1,558 | 19.7% (18.0%, 21.6%) | 9,235 | 17.2% (16.4%, 18.0%) |

<sup>1</sup>Population-weighted n. N represents the sample counts. The % values are based on weighted numbers.
<sup>2</sup>Population-weighted proportions and 95% confidence intervals (95% CI)

**Table 3:** Comorbidities prevalences distributed in binge drinking behavior and no binge drinking behavior groups.

| Characteristic | Overall |  | Binge drinking behavior |  | No binge drinking behavior |  |
| --- | --- | --- | --- | --- | --- | --- |
|  | N = 60,202<sup>1</sup> | % (95% CI)<sup>2</sup> | N = 8,104<sup>1</sup> | % (95% CI)<sup>2</sup> | N = 52,098<sup>1</sup> | % (95% CI)<sup>2</sup> |
| **Hypertension** |  |  |  |  |  |  |
| No | 47,702 | 78.6% (78.0%, 79.2%) | 6,978 | 86.3% (85.0%, 87.5%) | 40,724 | 77.4% (76.7%, 78.1%) |
|  | N = 60,202<sup>1</sup> | % (95% CI)<sup>2</sup> | N = 8,104<sup>1</sup> | % (95% CI)<sup>2</sup> | N = 52,098<sup>1</sup> | % (95% CI)<sup>2</sup> |
| Yes | 12,500 | 21.4% (20.8%, 22.0%) | 1,126 | 13.7% (12.5%, 15.0%) | 11,374 | 22.6% (21.9%, 23.3%) |
| **Type 2 diabetes** |  |  |  |  |  |  |
| No | 56,566 | 93.8% (93.4%, 94.1%) | 7,874 | 97.3% (96.7%, 97.8%) | 48,692 | 93.2% (92.8%, 93.6%) |
| Yes | 3,636 | 6.2% (5.9%, 6.6%) | 230 | 2.7% (2.2%, 3.3%) | 3,406 | 6.8% (6.4%, 7.2%) |
| **Heart problems** |  |  |  |  |  |  |
| No | 57,969 | 95.8% (95.5%, 96.1%) | 7,949 | 98.1% (97.6%, 98.5%) | 50,020 | 95.5% (95.1%, 95.8%) |
| Yes | 2,233 | 4.2% (3.9%, 4.5%) | 155 | 1.9% (1.5%, 2.4%) | 2,078 | 4.5% (4.2%, 4.9%) |
| **Stroke** |  |  |  |  |  |  |
| No | 59,236 | 98.5% (98.3%, 98.6%) | 8,058 | 99.3% (98.8%, 99.6%) | 51,178 | 98.3% (98.1%, 98.5%) |
| Yes | 966 | 1.5% (1.4%, 1.7%) | 46 | 0.7% (0.4%, 1.2%) | 920 | 1.7% (1.5%, 1.9%) |
| **Asthma** |  |  |  |  |  |  |
| No | 57,582 | 95.6% (95.3%, 95.9%) | 7,767 | 96.1% (95.3%, 96.7%) | 49,815 | 95.5% (95.2%, 95.8%) |
| Yes | 2,620 | 4.4% (4.1%, 4.7%) | 337 | 3.9% (3.3%, 4.7%) | 2,283 | 4.5% (4.2%, 4.8%) |
| **Arthritis/Rheumatism** |  |  |  |  |  |  |
| No | 56,226 | 93.6% (93.2%, 93.9%) | 7,864 | 97.1% (96.4%, 97.7%) | 48,362 | 93.0% (92.6%, 93.4%) |
| Yes | 3,976 | 6.4% (6.1%, 6.8%) | 240 | 2.9% (2.3%, 3.6%) | 3,736 | 7.0% (6.6%, 7.4%) |
| **Chronic spinal problem** |  |  |  |  |  |  |
| No | 49,624 | 81.5% (80.9%, 82.2%) | 6,881 | 85.3% (83.9%, 86.6%) | 42,743 | 80.9% (80.2%, 81.6%) |
| Yes | 10,578 | 18.5% (17.8%, 19.1%) | 1,223 | 14.7% (13.4%, 16.1%) | 9,355 | 19.1% (18.4%, 19.8%) |
| **WMSD** |  |  |  |  |  |  |
|  | N = 60,202<sup>1</sup> | % (95% CI)<sup>2</sup> | N = 8,104<sup>1</sup> | % (95% CI)<sup>2</sup> | N = 52,098<sup>1</sup> | % (95% CI)<sup>2</sup> |
| No | 59,053 | 97.6% (97.3%, 97.8%) | 7,943 | 97.3% (96.6%, 98.0%) | 51,110 | 97.6% (97.3%, 97.8%) |
| Yes | 1,149 | 2.4% (2.2%, 2.7%) | 161 | 2.7% (2.0%, 3.4%) | 988 | 2.4% (2.2%, 2.7%) |
| **Major depressive disorder** |  |  |  |  |  |  |
| No | 55,967 | 92.4% (91.9%, 92.8%) | 7,723 | 95.2% (94.4%, 95.9%) | 48,244 | 91.9% (91.4%, 92.4%) |
| Yes | 4,235 | 7.6% (7.2%, 8.1%) | 381 | 4.8% (4.1%, 5.6%) | 3,854 | 8.1% (7.6%, 8.6%) |
| **Other mental disorders** |  |  |  |  |  |  |
| No | 59,650 | 99.1% (98.9%, 99.2%) | 8,064 | 99.6% (99.4%, 99.8%) | 51,586 | 99.0% (98.8%, 99.1%) |
| Yes | 552 | 0.9% (0.8%, 1.1%) | 40 | 0.4% (0.2%, 0.6%) | 512 | 1.0% (0.9%, 1.2%) |
| **Lung problems** |  |  |  |  |  |  |
| No | 59,268 | 98.2% (98.0%, 98.4%) | 7,993 | 98.6% (98.1%, 99.0%) | 51,275 | 98.2% (97.9%, 98.4%) |
| Yes | 934 | 1.8% (1.6%, 2.0%) | 111 | 1.4% (1.0%, 1.9%) | 823 | 1.8% (1.6%, 2.1%) |
| **Cancer** |  |  |  |  |  |  |
| No | 59,179 | 98.2% (98.0%, 98.4%) | 8,054 | 99.2% (98.7%, 99.5%) | 51,125 | 98.0% (97.8%, 98.2%) |
| Yes | 1,023 | 1.8% (1.6%, 2.0%) | 50 | 0.8% (0.5%, 1.3%) | 973 | 2.0% (1.8%, 2.2%) |
| **Chronic kidney failure** |  |  |  |  |  |  |
| No | 59,363 | 98.6% (98.4%, 98.7%) | 8,028 | 99.1% (98.8%, 99.4%) | 51,335 | 98.5% (98.3%, 98.7%) |
| Yes | 839 | 1.4% (1.3%, 1.6%) | 76 | 0.9% (0.6%, 1.2%) | 763 | 1.5% (1.3%, 1.7%) |

<sup>1</sup>Population-weighted n. N represents the sample counts. The % values are based on weighted numbers.
<sup>2</sup>Population-weighted proportions and 95% confidence intervals (95% CI).

In the network analysis of binge drinking and comorbidities together, community 1 was represented by hypertension, type 2 diabetes, heart problems, stroke, and cancer, community 2 by asthma and lung problems, community 3 by arthritis/ rheumatism, chronic spinal problems, work-related musculoskeletal disorder (WMSD), and chronic kidney failure, and community 4 by major depressive disorder, and other mental disorders (Fig 1). Among all these communities, alcohol use was negatively related to almost all community 1 diseases and to arthritis/rheumatism in community 3. Looking at expected influence, binge drinking behavior negatively influenced the network structure.

**Fig 1.**
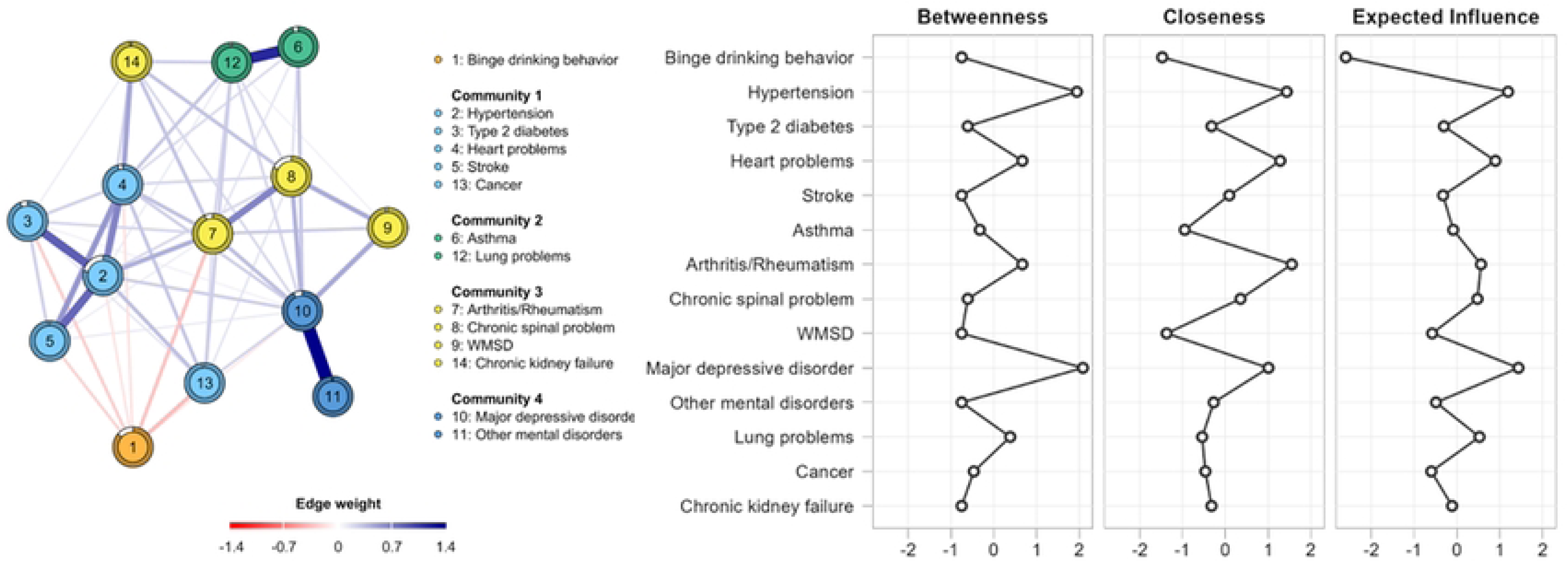
Binge drinking network and betweenness, closeness, and expected influence among comorbidities.

When comparing the comorbidities within binge drinking and no binge drinking behavior groups, networks were less connected in the binge drinking group and community connections changed between networks (Fig 2). Lung problems showed stronger connections with other mental disorders and major depressive disorders and became part of a new community in the binge drinking group composed of asthma, WMSD, major depressive episodes, other mental disorders, and lung problems. Cancer did not connect to other nodes in the binge drinking behavior network. Also, heart problems seemed to be more related to chronic kidney failure and arthritis/rheumatism, generating a new community including these nodes and chronic spinal problems.

**Fig 2.**
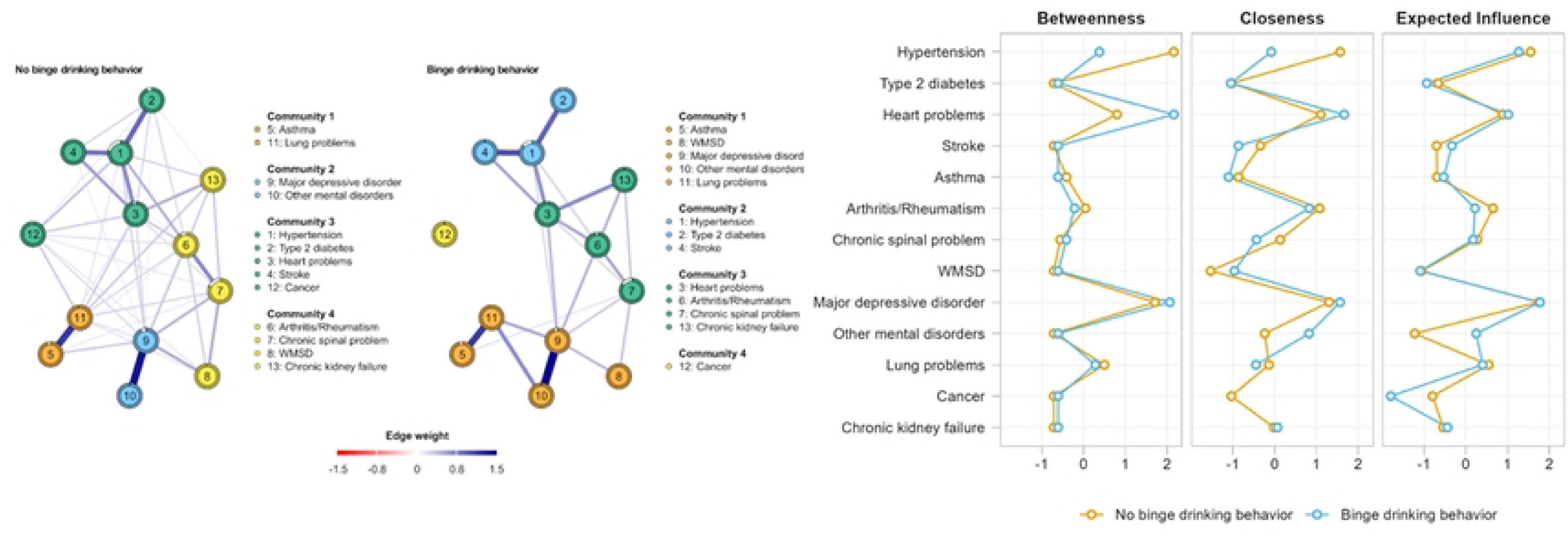
Networking and betweenness, closeness, and expected influence comparing different comorbidities within binge drinking and no binge drinking behavior groups.

The betweenness of hypertension was higher for the no binge drinking behavior network, while the variable heart problems had a higher betweenness for the binge drinking behavior network. In terms of expected influence, other mental disorders showed a higher value for the binge drinking behavior network than the other network. The closeness among binge drinking and no binge drinking behavior was very similar except for the higher values of hypertension closeness for the no binge drinking group (Fig 2).

Concerning the network analysis comparing different demographic characteristics, binge drinking was more strongly and negatively connected with stroke and cancer in the male than in the female network (S1 Figure, S2 Table). The male network showed stronger connections between nodes from Community 1 and other nodes of the network. Lung problems showed higher betweenness in the male network, while expected influence showed very similar patterns across the networks. Analyzing the network patterns through different ages, weak and negative edges were found between binge drinking behavior and major depressive disorder in people aged 25 to 44 years (S2 Figure, S2 Table). A weak negative connection between binge drinking and Arthritis/Rheumatism or major depressive disorder was also identified in the 45 to 64-year-old population. A sparser network was found for the 18 to 24-year network, revealing a low connection between morbidities in this age group. Binge drinking behavior stratified by age showed low scores for all centrality measures in the four networks. Major depressive disorder showed higher values of Expected Influence in all the networks.

Relating to race, binge drinking behavior showed stronger and negative edges with cancer, type 2 diabetes, and arthritis/rheumatism in the white network when compared to the black/parda network (S3 Figure, S3 Table). No connection was found between WMSD and lung problems in the white race network, unlike the results of the other networks. For the different regions, binge drinking behavior showed the strongest connection with stroke in the Northeast network when compared across all networks (S4 Figure, S4 Table). In addition, binge drinking behavior was not associated with other nodes in the post-secondary/equivalent network (S5 Figure, S5 Table). Lower levels of education showed more connections between binge drinking behavior and chronic conditions.

## Discussion

Using network analysis, our study aimed to understand the associations between binge drinking behaviors and the presence of co-morbid diseases across multiple socio-demographic dimensions. In general, we observed a higher prevalence of binge drinking in men in the 25-44 years age group, who identified as black/pardo, and those with secondary schooling or equivalent. The main network model revealed hypertension and chronic spinal problems as the main comorbidities among people with binge drinking. However, variations in the patterns of comorbidities across population groups belonging to different socio-demographic strata are noteworthy.

The overall prevalence of binge drinking in the studied population was found to be 13.5%, which is lower than the global prevalence for individuals over 15 years old in 2016 (18.2%), as reported by the World Health Organization (1). However, it should be noted that the exclusion of adolescents from the study may have influenced this result as the average age of first-time alcohol use among Brazilian adolescents is 13 years old (35). Furthermore, in our study, people aged 25-44 years old were the majority, while young adults represented 20.1% of that population. In terms of gender, there was an increase in weekly alcohol consumption among women from 2013 (23.9%) to 2019 (26%), according to the Brazilian Institute of Geographic and Statistics (36). Notably, the present study reports a prevalence of binge drinking in the women population of 25.5%, higher than the global heavy episodic drinking prevalence of 19.9% for this gender (1). This result can indicate an increased risk for binge drinking among women from Brazil, highlighting the need for attention toward this specific group. While the findings of the 2013 Brazilian National Health Survey, including alcohol use prevalence, have been described previously by Macinko et al (37), the current study focused on highlighting the extent of binge drinking behavior, variations in it across socio-demographic strata, and its association with various comorbidities. In this sense, we observed that the network of the binge drinking behavior group presented much fewer connections than the no binge drinking behavior network. This finding may be indicative of the influence that alcohol use exerts on the connections between multiple diseases. A less connected network suggests that alcohol use may positively impact the presence of certain conditions, such as hypertension or stroke, to the point that it weakens other associations. Hereupon, binge drinking is a recognized risk factor for several conditions, including cardiovascular diseases, type 2 diabetes, and arthritis/ rheumatism, among others (8). Additionally, our results may also reflect lower rates of binge drinking behavior among the elderly population, who are known to have a higher prevalence of comorbidities. In this regard, a study including adults over the age of 50 years with 2 or more comorbidities demonstrated that non-binge drinkers had a higher prevalence of multiple diseases compared to the binge-drinking group within that population (38).

The contrast between the network of individuals exhibiting binge drinking behavior and the network of non-binge drinkers is also evident in the patterns of connections within different disease communities. For instance, while community 3 within the no-binge drinking behavior group includes hypertension, type 2 diabetes, heart problems, stroke, and cancer, cancer represents its own community on the binge drinking behavior network. This disparity suggests that alcohol is altering the patterns of connections between comorbidities, possibly exerting a substantial effect on the development of cancer. In fact, alcohol is a well-established risk factor for cancer, including gastric, colorectal, pharyngeal, and breast cancer, and surpasses other potential risk factors in its influence (8). Furthermore, even after diagnosis, alcohol consumption remains a common habit among individuals with cancer. A survey conducted in the United States revealed that among participants diagnosed with cancer over five years before the study, 57.1% reported being current drinkers, with 20.1% engaging in binge drinking behavior (10).

Our analysis of the correlation between binge drinking and comorbidities within different socio-demographic variables highlighted distinct connections for each group. For example, although it is stated in the literature a positive correlation between alcohol use and mental and behavioral diseases(8,11,39), our findings pointed to a weak negative correlation between binge drink and major depressive disorder (MDD) in patients aged 25 to 64 years old. In addition, the difference between sexes pointed to a strongly negative correlation between binge drink and cancer in males, compared to females. Despite that, there is a known correlation between alcohol and cancer, including cancers more frequent in men, such as colorectal, esophagus, throat, and liver cancers (40). About different ethnicities, the white population presented a stronger negative correlation between binge drinking and cancer, type 2 diabetes, and arthritis/ rheumatism. Moreover, although a negative correlation between higher education level and heavy drinkers is well known (41), stronger negative associations between binge drinking and chronic comorbidities were present in the lower educational levels group.

In the last two decades, network models have gained rapid popularity in health-related analyses to identify patterns of multimorbidity. Through multivariate modeling and sensitivity analyses, the application of networks attempts to understand population-level patterns of multimorbidity associations. These models have been used across different contexts and populations such as elderly health (13), cerebrovascular diseases (12), heart failure, and chronic obstructive pulmonary disease (42), as well as the correlation of multimorbidities and risk of COVID-19 hospitalization and mortality (43). Furthermore, the easy visualization of information allows the identification of patterns and, subsequently, the development of studies and the practical application of the findings (44). The present study used network analysis to identify patterns of comorbidities in binge-drinking populations in Brazil.

### Strengths and Limitations

Although network models can give insights into the causal pathways between nodes, directionality detection is not possible. Since some of the chronic conditions included in the study could result in death, this study naturally failed in the recruitment of potential participants who may not have survived until the date of the data collection. In addition, recall bias may have affected the results, given that all the measures were self-reported, exposing the data to possible information misreporting situations and inherent bias. Our study is also limited by the small binge-drinking population sample size. It is important to note that this study used the Brazilian National Survey from 2013 and some of these findings may be outdated, mainly, it does not reflect the impacts of the COVID-19 pandemic.

Despite the cited limitations, our study descriptively demonstrated the existence of groups more prone to binge drinking behavior. The use of network analysis allows a visual interpretation of the relevant variables in the context of the occurrence of health problems. This methodology enables an interpretation of the perspectives related to comorbidities, demonstrating elements that are common to all groups analyzed and allowing the formulation of hypotheses about the nature of comorbidities and, respectively, the implementation of specific policies and treatments based on the reality of the community (45). Furthermore, it is important to emphasize that the chances of error occurrence are considerably lower due to the extensive tests performed so that they can be conducted in an automated manner (46).

## Data Availability

Data is public available at https://ghdx.healthdata.org/record/brazil-national-survey-health-2013

## Authors’ contributions

SZ, DF, and JRNV performed the data curation, formal analysis, and methodology. PCPS and NDP participated in the original draft writing and review. SD CAS and JRNV contributed to conceptualization, project administration, and writing review. All the authors reviewed and accepted the final version.

## Ethics Statement

All data used for this analysis was obtained through a national health survey. Thus, this secondary data analysis used only de-identified publicly available data. Participants were not directly consented by the research team since all data was collected as a national policy survey. Data and documentation on the data set and data collection procedures are available at: https://www.pns.icict.fiocruz.br/

## Supplementary data

S1 Table. Chronic conditions and binge drinking prevalences according to sex.

S1 Figure. Chronic conditions and binge drinking Network according to sex.

S2 Table. Chronic conditions and binge drinking prevalences according to age.

S2 Figure. Chronic conditions and binge drinking Network according to age.

S3 Table. Chronic conditions and binge drinking prevalences according to ethnicity.

S3 Figure. Chronic conditions and binge drinking Network according to ethnicity.

S4 Table. Chronic conditions and binge drinking prevalences according to region.

S4 Figure. Chronic conditions and binge drinking Network according to region.

S5 Table. Chronic conditions and binge drinking prevalences according to educational level.

S5 Figure. Chronic conditions and binge drinking Network according to educational level.

